# Revisiting the Role of Structural Connectivity-Based Parcellation in Thalamic Nuclei Segmentation: comparison with recent state-of-the-art methods

**DOI:** 10.1101/2025.09.16.25335779

**Authors:** Daniel H. Nguyen, Vinod Kumar, Debottama Das, Ali Bilgin, Dianne Patterson, Alberto Cacciola, Manojkumar Saranathan

**Author notes:** Corresponding author: Manojkumar Saranathan, PhD. Full postal address: 55 Lake Avenue North, Worcester, MA, 01655.

## Abstract

Accurate thalamic nuclei segmentation is critical for neuroscience research and clinical interventions such as deep brain stimulation and magnetic resonance guided focused ultrasound. Connectivity based parcellation has been widely used for two decades, yet its anatomical validity remains uncertain compared with newer imaging approaches.

**Methods:** We analyzed high resolution diffusion magnetic resonance imaging (MRI) and T1 weighted data from 67 healthy young adults in the Human Connectome Project. Connectivity based parcellation was performed using probabilistic tractography with cortical targets derived from the HCP MMP1 atlas, generating 8, 11, and 23 region parcellations. Results were compared against three state of the art methods: orientation distribution function (ODF) clustering, track density imaging (TDI), and the structural MRI based segmentation. Group level analyses were conducted in Montreal Neurological Institute and Hospital (MNI) space, and Dice overlap coefficients were calculated against the histology based Morel atlas.

**Results:** Connectivity based parcellation demonstrated limited anatomical precision, with increasing cortical target counts introducing greater variability and noise without improving nuclear boundary definition. ODF clustering and TDI recovered subdivisions consistent with cytoarchitectonic patterns, particularly in the pulvinar and mediodorsal nuclei. Structural MRI based segmentation achieved the highest overall Dice coefficients, closely approximating Morel defined boundaries, while Connectivity based parcellation consistently underperformed across nuclei.

**Conclusion:** Despite methodological advances, Connectivity based parcellation remains constrained in its ability to delineate thalamic nuclei with histological accuracy. By contrast, structural and diffusion microstructural (ODF, TDI) approaches provide superior nuclear localization. These findings highlight the need for hybrid workflows that integrate structural and diffusion based information to enable more reliable thalamic segmentation for research and clinical targeting applications.

## Introduction

The thalamus is a subcortical brain structure that plays a central role in relaying and integrating sensory, motor, and cognitive information between cortical, subcortical, and cerebellar regions. It is functionally organized into individual thalamic nuclei, each characterized by unique connectivity patterns to specific brain regions. These nuclei are highly relevant in a variety of diseases, with structural and functional alterations often linked to specific clinical scenarios. For instance, the ventral posterolateral (VPL) nucleus of the thalamus is relays somatosensory information^1^ and has been associated with neuropathic pain and nociception^2^, whereas the mediodorsal (MD) nucleus is associated with executive function and has been implicated in chronic neurodegenerative diseases, including frontotemporal dementia^3^. The pulvinar (Pul) nucleus, its widespread connections to visual, auditory, somatosensory, parietal, temporal, and occipital cortices, plays a critical role in higher-order sensory integration and multimodal processing ^4–7^. Dysfunction of the mesial pulvinar has been linked to temporal lobe epilepsy, where seizures propagate through its extensive cortical connections ^8^. Given their functional importance, thalamic nuclei have also emerged as therapeutic targets. Deep brain stimulation (DBS), a neuromodulatory treatment that delivers electrical stimulation to specific brain regions, has traditionally been used to treat tremor in Parkinson’s disease (PD) by targeting the subthalamic nucleus ^9,10^, but has also been applied to thalamic nuclei for drug-resistant epilepsy ^11^ and several movement disorders, notably essential tremor ^12,13^. Similarly, Magnetic Resonance–guided Focused UltraSound (MRgFUS) enables lesioning of specific nuclei such as the ventral intermediate (Vim) nucleus of the thalamus, for the treatment of essential tremor ^13^ and tremor dominant PD ^14,15^. These interventions critically rely on accurate localization of desired nuclei, traditionally guided by a combination of standardized stereotactic coordinates, anatomical landmarks and/or by cytoarchitectonic atlases such as the Schaltenbrand Atlas^16^, or, the Morel thalamic atlas ^17^. However, atlas-based targeting has notable limitations, including reduced reliability/consistency in nucleus boundaries across patients ^18^.

Over the past two decades, several methods for direct and precise delineation of thalamic nuclei have been proposed. Connectivity-based parcellation (CBP) emerged with the advent of diffusion-weighted imaging (DWI), leveraging connectivity patterns to assign thalamic voxels to specific cortical regions of the brain^19^. Behrens et. al. (2003) pioneered this approach, segmenting the thalamus into seven different cortical connectivity-based parcels ^19^. Since the seminal work of Behrens et al. (2003), other studies have confirmed the reproducibility of connectivity-based parcellation (CBP) while refining its methodology^20^. For instance, a 2010 study demonstrated that thalamic CBP is highly reproducible when using broad cortical targets, but reproducibility decreases substantially with finer parcellations involving 31 cortical targets^21^. CBP has also been extended to the subthalamic region ^22^, with another study showing distinct subdivisions of the MD nucleus that align well with known functional and anatomical territories^23^.

Other approaches based on diffusion MRI at a local level without requiring cortical target definitions have been proposed. The most successful of them use diffusion tensor imaging (DTI)^24^ and involve different ways to cluster the primary angular orientation of the diffusion tensor ^25–27^. A more recent study used spherical harmonic representations of orientation distribution function (ODFs) to differentiate intra-thalamic microstructures ^28^. ODF-based parcellation was demonstrated to be much more stable and reproducible compared to angular direction approaches. Basile et al. (2021) introduced short-tracks track-density imaging (TDI) providing maps with similarities to that of a histological thalamic atlas ^29^. The utilization of resting-state functional magnetic resonance imaging (fMRI) emerged shortly after the introduction of CBP, identifying functional subdivisions of the thalamus through temporal correlations in spontaneous BOLD signal fluctuations between thalamic voxels and the cortical network^30^. Early work by Zhang et al. (2008) demonstrated strong functional specialization of individual thalamic voxels^31^, which was later refined by using temporally independent thalamocortical states^30^ and instantaneous connectivity parcellation to delineate functionally distinct subregions^32^.

The primary reason for the dominance of diffusion, and to an extent, fMRI-based methods for thalamic nuclei segmentation until recently was the poor intrathalamic contrast of structural T1 and T2 weighted MRI. In the last 5 years, several advances have resulted in the resurgence of structural MRI based thalamic nuclei segmentation. A probabilistic atlas-based Bayesian segmentation from ex-vivo histology and in-vivo Magnetization Prepared Rapid Gradient Recalled Echo (MP-RAGE) T1 weighted MRI has been proposed and is part of Freesurfer^33^.

Specialized sequences such as Fast Gray Matter Acquisition T1 Inversion Recovery (FGATIR) ^34^ have been proposed for improved visualization of thalamic nuclei relative to standard T1- and T2-weighted MRI ^35,36^. These have gone hand-in-hand with segmentation methods such as Thalamus Optimized Multi-Atlas Segmentation (THOMAS) technique^37^ which leverages the improved contrast of white-matter-nulled MP-RAGE^38^ and its recent extension Histogram-based polynomial synthesis (HIPS)-THOMAS which synthesizes white-matter-nulled images from conventional T1weighted MRI ^39^. However, to our knowledge, no studies have comprehensively compared and evaluated CBP methods against other recent parcellation methods in the same cohort of subjects.

Thus, in this study, we sought to revisit CBP, leveraging state-of-the-art high resolution diffusion MRI data, advanced diffusion processing methods such as multi-shell multi-tissue constrained spherical deconvolution (MSMT-CSD) that account for tissue properties and handle crossing fibres, and cortical parcellations derived from sophisticated atlases such as the Glasser HCP- MMP1 atlas ^40^. Our first aim was to assess the performance of “modern” CBP, segmenting the thalamus into functional parcellations based on cortical connectivity, with the hopes of achieving parcellations that achieve closer resemblance to histological-based atlases. We then compared CBP-derived parcellations to three state-of-the-art methods leveraging diffusion microstructure, short-range structural connectivity, and structural MRI, to order to assess their relative anatomical validity and reproducibility.

## Materials & Methods

### Participants and Imaging Data

Data used in this study were sourced from the publicly available Human Connectome Project (HCP) Young Adult (YA) 1200 Subjects dataset ^41^. A subset of n = 67 healthy young adults (ages 22-35) was selected based on the availability of both structural T1-weighted and high- resolution diffusion MRI data. All participants provided informed consent, and the Institutional Review Board approved the HCP protocols.

### MRI Acquisition

A schematic of the entire workflow can be found in **Figure 1**. Structural T1-weighted images (HCP-YA) were acquired using a 3D MPRAGE sequence with 0.7 mm isotropic resolution (TR = 2400 ms, TE = 2.14 ms, TI = 1000 ms, flip angle = 8°) ^41^. High-resolution diffusion MRI data (HCP-YA) were acquired using a spin-echo EPI sequence with 1.25 mm isotropic resolution, sampling three shells (b = 1000, 2000, 3000 s/mm²) across 270 diffusion directions, plus 18 b=0 volumes. Acquisition was performed with both right-to-left and left-to-right phase encoding to enable correction for susceptibility-induced distortions ^42,43^.

**Figure 1:**
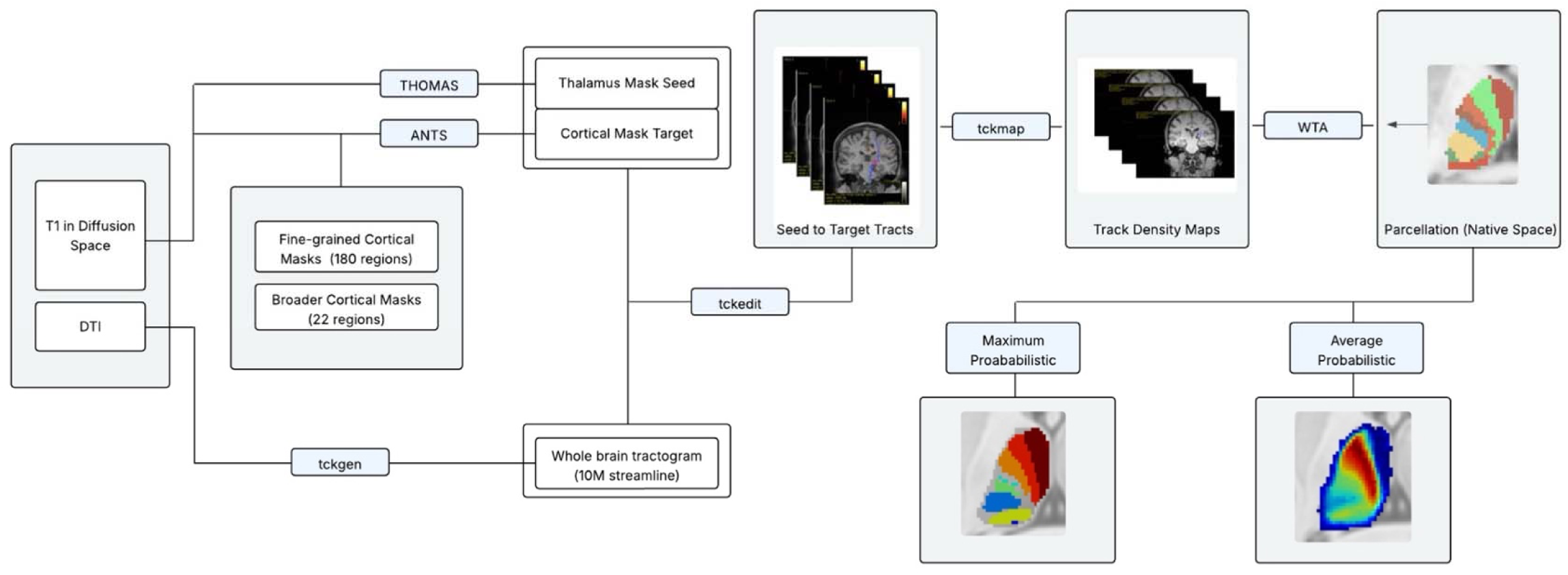
Workflow for Generating CBP of HCP Subjects. T1-weighted and diffusion MRI were processed using ANTs and MRtrix3. Thalamic nuclei were segmented with HIPS- THOMAS ^39^, cortical regions were defined using the HCP-MMP1 atlas ^40^, and probabilistic tractography was performed to assess connectivity between specific thalamic and cortical regions. Connectivity results were converted into track density maps, used for winner-take-all thalamic parcellations, and aggregated into group-level probability maps in MNI space.

### Image Preprocessing

Structural MRI images were preprocessed using standard methods, including skull-stripping and bias-field correction. T1-weighted images were subsequently coregistered to diffusion space using Advanced Normalization Tools (ANTs) affine registration. DWI data underwent standard preprocessing using MRtrix3, including denoising, eddy-current and motion correction, bias field correction, and normalization of diffusion-weighted signal intensity.

### Cortical Parcellation Schemes

Cortical regions were delineated using the volumetric version of the Glasser Human Connectome Project multimodal parcellation atlas (HCP-MMP1), which can be found at https://osf.io/azup8, comprising 180 cortical parcels per hemisphere called “areas”. From these 180 “areas”, these can be further grouped into 22 larger “regions”, each comprising different functional characteristics as described in Glasser et al ^40^. Subject-specific T1-weighted images, which have already been coregistered to diffusion space, were nonlinearly registered to the MNI152 (ICBM 2009a Nonlinear Asymmetric) template using ANTs (*antsRegistrationSyNQuick.sh*) with a symmetric normalization (SyN) transformation, producing both an affine matrix and nonlinear warp fields. These transformations were inverted to project the HCP-MMP1 atlases (22- and 180-region) into each subject’s native T1/diffusion space using *antsApplyTransforms* with nearest-neighbor interpolation, yielding subject-space cortical parcellations to be used for the cortical target ROI. The 22- and 180-region atlases were used to create 3 cortical parcellation schemes (henceforth referred to as 8-parcellation, 11-parcellation, and 23-parcellation). The 8-parcellation essentially follows the same cortical scheme as Behrens et al.^19^, with the addition of the frontal region. The 23-parcellation is derived from the 22-region atlas, with a split of the region that is defined as “Somatosensory and Motor Cortex” into separate somatosensory and motor cortical regions.

Finally, the 11-parcellation is a middle ground between the two parcellation schemes. Details of each of these parcellation schemes can be found in **Table 1** and visualized in **Figure S1**.

**Table 1:** Glasser Atlas Area Division for each Parcellation Scheme.

|  |  |
| --- | --- |
| 8-Parcellation | **Temporal:** EC, PreS, H, PeEc, PHA2, TF, TGd, TGv, PHA1, PHA3, TE1a, TE1p, TE2a, TE2p, TE1m, PHT, H_L<br>**Parietal:** PSL, STV, TPOJ1, TPOJ2, TPOJ3, 7Pm, 7AL, 7Am, 7Pl, 7PC, LIPv, VIP, MIP, AIP, LIPd, PFt, AIP, PFop, PF, PFm, PGi, PGs, PGp, IP2, IP1, IP0<br>**Motor:** 4<br>**Sensory:** 3b, 1, 2, 3a<br>**Visual:** V1, V2, V3, V4, V6, V3A, V7, V3B, IPS1, DVT, V6A, V8, PIT, FFC, VMV1, VMV2, VMV3, VVC, MST, LO1, LO2, MT, V4t, FST, V3CD, LO3, PH<br>**Premotor:** FEF, PEF, 55b, 6d, 6v, 6a, 6r<br>**Prefrontal:** 47m, 10pp, 10d, 10r, p10p, 47s, OFC, 13l, 11l, 44, 45, 47l, a47r, 6r, IFJa, IFJp, IFSp, IFSa, p47r, SFL, 8Av, 8Ad, 8BL, 9p, 8C, p9-46v, 46, a9-46v, 9-46d, 9a, i6-8, s6-8<br>**Frontal:** p24pr, 33pr, a24pr, p32pr, a24, d32, 8BM, p32, 10r, 9m, 10v, 25, s32, pOFC, a32pr, p24 |
| 11-Parcellation | **Visual:** V1<br>**Visual Streams:** V2, V3, V4, V6, V3A, V7, V3B, IPS1, DVT, V6A, V8, PIT, FFC, VMV1, VMV2, VMV3, VVC, MST, LO1, LO2, MT, V4t, FST, V3CD, LO3, PH, TPOJ3<br>**Motor:** 4<br>**Sensory:** 3b, 1, 2, 3a<br>**Cingulate Tract:** 5m, 5mv, 23c, 5L, 24dd, 24dv, SCEF, 6mp, 6ma, RSC, POS2, PCV, 7m, POS1, 23d, v23ab, d23ab, 31pv, ProS, DVT, 31pd, 31a, p24pr, 33pr, a24pr, p32pr, a24, d32, 8BM, p32, 10r, 9m, 10v, 25, s32, pOFC, a32pr, p24<br>**Premotor:** FEF, PEF, 55b, 6d, 6v, 6a, 6r<br>**Posterior Opercular:** 43, OP4, OP1, OP2-3, FOP1<br>**Prefrontal:** 47m, 10pp, 10d, 10r, p10p, 47s, OFC, 13l, 11l, 44, 45, 47l, a47r, 6r, IFJa, IFJp, IFSp, IFSa, p47r, SFL, 8Av, 8Ad, 8BL, 9p, 8C, p9-46v, 46, a9-46v, 9-46d, 9a, i6-8, s6-8<br>**Parietal:** PSL, STV, TPOJ1, TPOJ2, TPOJ3, 7Pm, 7AL, 7Am, 7Pl, 7PC, LIPv, VIP, MIP, AIP, LIPd, PFt, AIP, PFop, PF, PFm, PGi, PGs, PGp, IP2, IP1, IP0, PSL, STV, TPOJ2<br>**Temporal:** EC, PreS, H, PeEc, PHA2, TF, TGd, TGv, PHA1, PHA3, TE1a, TE1p, TE2a, TE2p, TE1m, PHT, TPOJ1<br>**Auditory:** A1, 52, RI, PFcm, PBelt, MBelt, LBelt, TA2, STGa, A5, STSda, STSdp, STSvp, STSva, A4, PoI2, MI, Pir, AVI, AAIC, FOP3, FOP2, FOP4, FOP5, PI, PoI1, Ig |
| 23-Parcellation | **Primary Visual:** V1<br>**Early Visual:** V2, V3, V4<br>**Dorsal Stream Visual:** V6, V3A, V7, V3B, IPS1, DVT, V6A<br>**Ventral Stream Visual:** V8, PIT, FFC, VMV1, VMV2, VMV3, VVC<br>**MT+ Complex and Neighboring Visual Areas:** MST, LO1, LO2, MT, V4t, FST, V3CD, LO3, PH<br>**Motor:** 4<br>**Somatosensory:** 3b, 1, 2, 3a<br>**Premotor:** FEF, PEF, 55b, 6d, 6v, 6a, 6r<br>**Posterior Cingulate:** RSC, POS2, PCV, 7m, POS1, 23d, v23ab, d23ab, 31pv, ProS, DVT, 31pd, 31a<br>**Paracentral Lobular and Mid Cingulate:** 5m, 5mv, 23c, 5L, 24dd, 24dv, SCEF, 6mp, 6ma<br>**Superior Parietal:** 7Pm, 7AL, 7Am, 7Pl, 7PC, LIPv, VIP, MIP, AIP, LIPd<br>**Anterior Cingulate and Medial Prefrontal:** p24pr, 33pr, a24pr, p32pr, a24, d32, 8BM, p32, 10r, 9m, 10v, 25, s32, pOFC, a32pr, p24<br>**Dorsolateral Prefrontal:** SFL, 8Av, 8Ad, 8BL, 9p, 8C, p9-46v, 46, a9-46v, 9-46d, 9a, i6-8, s6-8<br>**Inferior Frontal:** 44, 45, 47l, a47r, 6r, IFJa, IFJp, IFSp, IFSa, p47r<br>**Orbital and Polar Frontal:** 47m, 10pp, 10d, 10r, p10p, 47s, OFC, 13l, 11l<br>**Posterior Opercular:** 43, OP4, OP1, OP2-3, FOP1<br>**Early Auditory:** A1, 52, RI, PFcm, PBelt, MBelt, LBelt<br>**Auditory Association:** TA2, STGa, A5, STSda, STSdp, STSvp, STSva, A4<br>**Insular and Frontal Opercular:** PoI2, MI, Pir, AVI, AAIC, FOP3, FOP2, FOP4, FOP5, PI, PoI1, Ig<br>**Medial Temporal:** EC, PreS, H, PeEc, PHA2, TF, TGd, TGv, PHA1, PHA3<br>**Lateral Temporal:** TE1a, TE1p, TE2a, TE2p, TE1m, PHT<br>**Temporo-Parieto-Occipital Junction:** PSL, STV, TPOJ1, TPOJ2, TPOJ3<br>**Inferior Parietal:** PFt, AIP, PFop, PF, PFm, PGi, PGs, PGp, IP2, IP1, IP0 |

### Connectivity-based parcellation

Probabilistic tractography was conducted using *MRtrix3* software to estimate structural connectivity between the whole thalamus and cortical regions. Whole brain tractograms consisting of ten million streamlines per subject were generated for each of the n = 67 HCP subjects. Streamlines connecting the *whole* thalamus (segmented using HIPS-THOMAS, see description below) and specific cortical regions (defined by HCP-MMP1) were filtered using MRtrix’s *tckedit* function with default parameters, employing each of the masks as inclusion regions of interest (ROIs). The following connectivity analyses were performed: Whole thalamus connectivity to global cortical parcellations (8-parcellation, 11-parcellation, and 23- parcellation). Track density maps (TDMs) were computed from the filtered streamline datasets. Each TDM was normalized by using the thalamus mask as a spatial reference template to ensure consistent comparisons across subjects. A Winner-Take-All (WTA) approach was applied to assign each voxel within the thalamic nuclei to the cortical region demonstrating the highest normalized connectivity. This resulted in subject-specific, connectivity-driven thalamic segmentation maps (parcellations) in native space, one for each of the three parcellation schemes.

### Comparative Analyses

To critically compare CBP with state-of-the-art methods for thalamic nuclei segmentation, comparative analyses were conducted between CBP-derived segmentations, specifically the 8- parcellation, to ODF-based k-means clustering, TDI-based parcellation, as well as HIPS-THOMAS structural MRI segmentation. All results were visualized using standardized slice orientations in MNI space and consistent color-coding schemes were used as much as possible to facilitate direct comparison across methods. Quantitative metrics for overlap (Dice coefficients) were also computed for the four methods with the Morel Atlas serving as the ground truth. The resulting scores were summarized and displayed in heatmap form. The three methods used for comparisons are briefly described below.

#### ODF-Clustering

Thalamic parcellation using ODF clustering was performed following the framework described by Battistella et al. (2017) ^28^. Briefly, diffusion MRI data were first modeled using a spherical harmonic (SH) representation of the orientation distribution functions to provide full angular characterization of the diffusion process within each voxel. K-means clustering, initialized in a data-driven manner, was then applied to group thalamic voxels using a weighted average of Euclidean distance of voxel positions and SH coefficients as a distance metric, yielding seven reproducible thalamic nuclear clusters. The whole thalamus mask from HIPS-THOMAS was used instead of Freesurfer as in Battistella et al. (2017) but hyperparameters were kept identical.

#### Track Density Imaging

A probabilistic version of a TDI thalamic atlas based on manual parcellation of individual TDI from 6 HCP subjects (3 males, 3 females) ^29^ was used, freely available at https://github.com/BrainMappingLab/TDI-derived-Thalamic-Atlas. Briefly, for each subject, a manually defined bounding-box ROI encompassing the diencephalon was drawn using anatomical landmarks to optimize streamline density and TDI contrast. Within this ROI, short-tract tractograms were generated using the iFOD2 algorithm and from these tractograms, two super-resolution short-track TDI (stTDI) maps (0.25 mm isotropic) were produced: (i) a directionally encoded color stTDI (DEC-stTDI) and (ii) an apparent fiber density–weighted stTDI (AFD-stTDI). Manual delineation of 13 thalamic nuclei (anterior, centromedian-parafascicular, habenula, lateral dorsal, lateral geniculate, mediodorsal-centrolateral, mediodorsal, medial geniculate, midline nuclei, pulvinar, ventral anterior, ventral lateral and ventral posterior) was performed on stTDI maps. Segmentation was performed in sagittal slices using DEC-stTDI maps (with AFD-stTDI overlays at ∼80% opacity), guided by histological sections ^44^. The resulting 3D volumes were refined in axial and coronal planes, median filtered, eroded to minimize boundary overlap, binarized, and resampled to 0.5 mm³ resolution.

### HIPS-THOMAS

For structural MRI based segmentation, HIPS-THOMAS ^39^, a variant of the original THOMAS ^37^ method that is optimized for T1 weighted MRI was used. It uses a simple polynomial algorithm to synthesize WMn-like images prior to THOMAS. THOMAS itself is a classic multi-atlas segmentation method that uses a multi-atlas of 20 WMn-MPRAGE datasets, each manually segmented using the Morel atlas as a guide. After warping them to native space using ANTs nonlinear registration, the 20 segmentations are combined into a single final segmentation using a joint label fusion algorithm which uses local image similarity to differentially weight the labels ^45^.

### Group-Level Analysis

Native space parcellations results were spatially transformed into a standardized MNI template (ICBM 2009a Nonlinear Asymmetric) space using ANTs nonlinear registration. These parcellations, now all in MNI space, were then analyzed across the n = 67 individuals (n = 6 for TDI) using three approaches:

**1.** Maximum Relative Probability Maps: To compute the maximum probability label at each voxel across subjects, a voxel-wise majority voting strategy was performed. In this approach, all subject parcellations (in MNI space) were stacked voxel-wise. For each voxel in the thalamus, the most frequently assigned parcel label across individuals was identified. This resulted in a maximum relative probability map in which each voxel was labeled according to the most common assignment across the cohort.
**2.** Average Probability Maps: After stacking the subject parcellations in MNI space voxel-wise, binary masks were generated for each label and averaged across subjects, yielding voxel-wise probability maps. Each probability map was multiplied by the corresponding label index, and the results were summed to create a single weighted image. This representation highlights dominant nuclei while also reflecting the degree of labeling uncertainty across subjects, which appears as fuzzy boundaries.
**3.** Thresholded Relative Probability Maps: In this approach, all subject parcellations (in MNI space) were stacked voxel-wise. At each voxel, the frequency of each nucleus label across subjects was computed, producing probability maps. The voxel was assigned to the label with the highest probability, provided that the winning label exceeded a pre-determined (e.g. 25%) threshold. Voxels that failed to meet this criterion were reset to background (0). This approach reduces noise by excluding low-probability voxels, retaining only the most frequently labeled regions across the cohort.

## Results

### Thalamus Connectivity with 8-Parcellation

CBP between the thalamus and eight broadly defined cortical regions of interest (**Figure S1A**) is visualized in **Figure 2A** as the relative maximum probability maps and **Figure 3A** as the average probability maps. The relative maximum probability maps depict each of the thalamic-cortical connections by a specific color, revealing the layered arrangement of parcels across axial and coronal slices. For comparison, the parcellation map from Behrens et al. (2003) is also displayed at 25% maximum probability in comparison to the 8-parcellation at 25% maximum probability, demonstrating a similar layering pattern **(Figure S2A, Figure S2D).** This similar pattern is also observed when comparing the parcellation map from Behrens et al. (2003) to the 8-parcellation at relative maximum probability (**Figure 2A, 2D**). In their parcellation, the prefrontal cortex was represented as a single region (light red), whereas in our 8-parcellation this region is subdivided into prefrontal (light red) and frontal (dark red) components. Consistent with Behrens et al., we observe a distinct anterior-posterior layering pattern on the axial slice: premotor, motor, sensory, parietal, and visual cortical, a pattern that is also recapitulated in our 8-parcellation (**Figure 2A**, **Figure S2A**).

**Figure 2:**
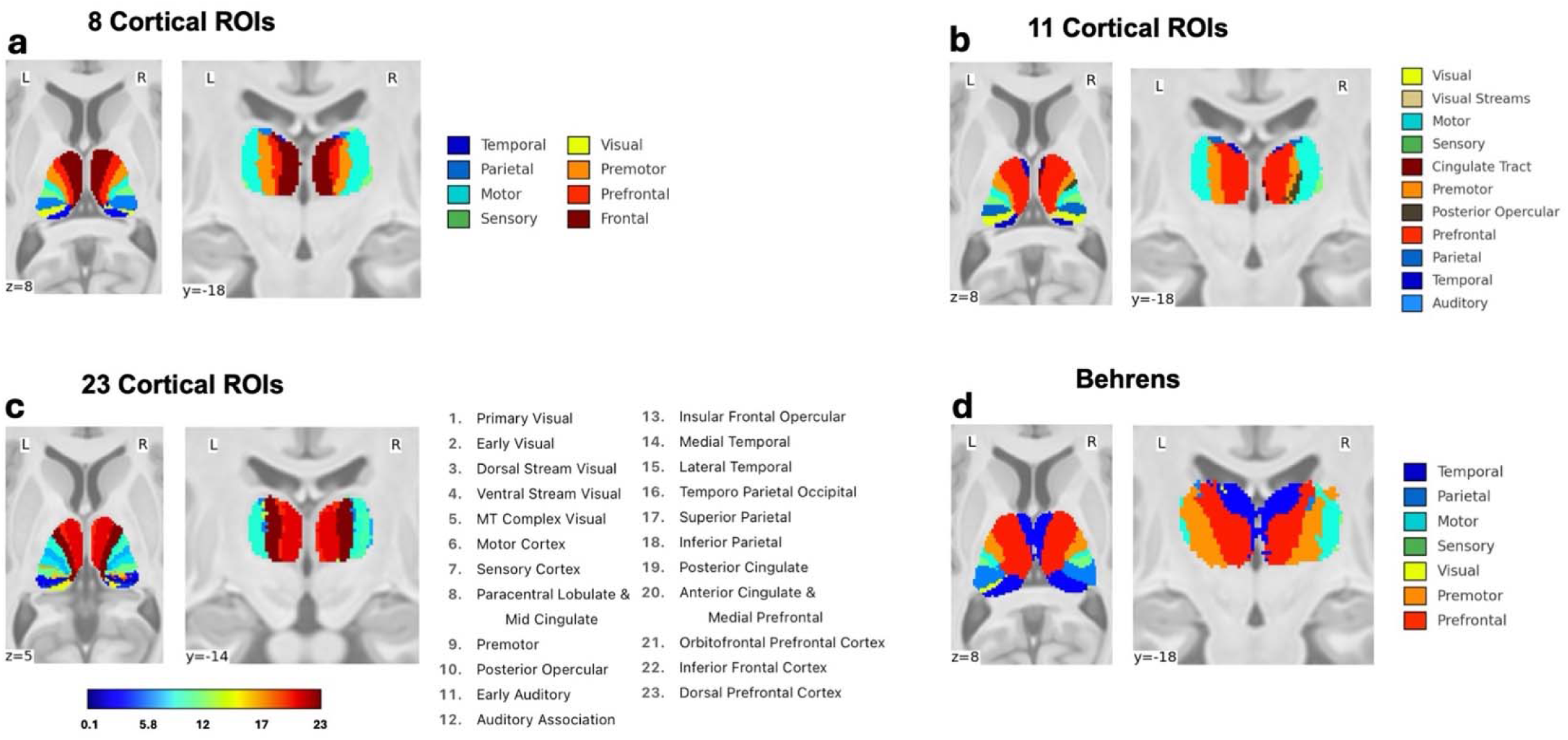
CBP between Thalamus and 8/11/23 Cortical Regions of Interest with Comparison to Behrens using Relative Maximum Probability. The following figure depicts cortical-based parcellation (CBP) between the thalamus mask (segmented using HIPS-THOMAS ^39^) and generating streamlines to 8 cortical regions referenced from Glasser et al (2016) ^40^. These regions were selected to be the 7 original cortical regions that Behrens et. al (2003) ^19^ had used, in addition to the Frontal Region (dark red). This figure also shows connections to 11- and 23- “broad” cortical regions referenced from Glasser et al (2016) ^40^. Behrens, visualized in the same MNI space, is also shown for comparison. Each parcellation scheme shows the axial view and the coronal view.

**Figure 3:**
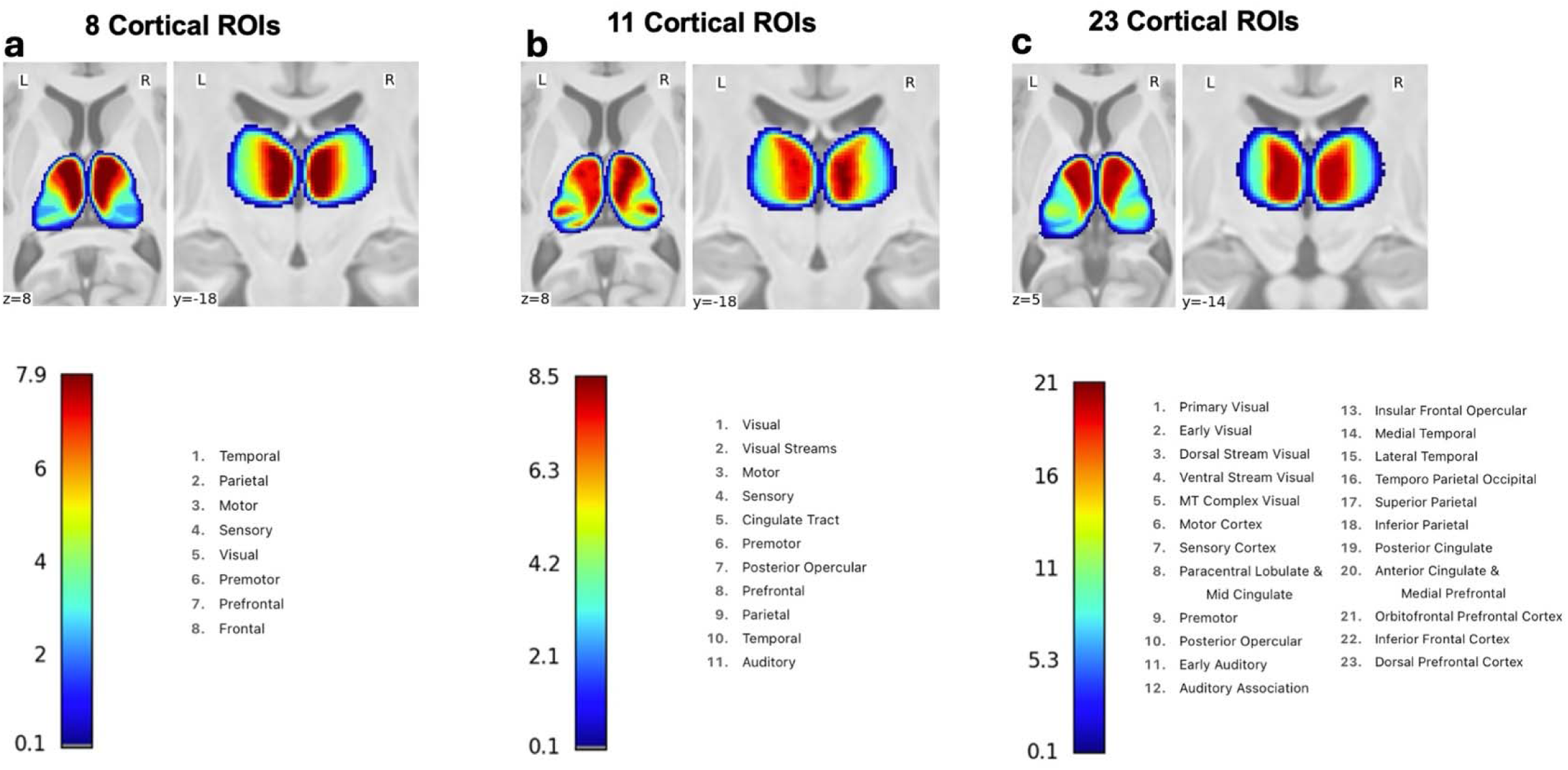
CBP between Thalamus and 8/11/23 Cortical Regions of Interest using Averaging. The following figure depicts cortical-based parcellation (CBP) between the thalamus mask (segmented using HIPS-THOMAS ^39^) and generating streamlines to 8 cortical regions referenced from Glasser et al (2016) ^40^. Voxels in each of the parcellations were averaged across n = 67 subjects to generate the image displayed. These regions were selected to be the 7 original cortical regions that Behrens et. al (2003) ^19^ had used, in addition to the Frontal Region (dark red). This figure also shows connections to 11- and 23- “broad” cortical regions referenced from Glasser et al (2016) ^40^. Each heatmap displays the ranges of Label IDs for each of the parcellation schemes, along with the corresponding cortical region for each Label ID. Each parcellation scheme shows the axial view and the coronal view.

### Thalamus Connectivity with 23-Parcellation

The volumetric HCP-MMP1 atlas is already grouped by 23 regions (**Figure S1C**), and CBP between the thalamus and these 23 regions (with the splitting of region six into its own, individual regions: sensory and motor) were explored with the aim of generating a more finely grained thalamus parcellation (**Figure 2C**, **Figure 3C**). However, we found that this parcellation scheme resulted in noisier parcellation patterns (e.g. auditory Label 11, **Figure 2C**, and posterior cingulate cortices Label 19, **Figure 2C**). Interestingly, after applying a 25% probability threshold, the posterior thalamic regions were largely absent (**Supplementary Figure 2C**), suggesting that these voxels lack consistent connectivity patterns across subjects. This variability points to poor reproducibility of posterior thalamic labels in the 23-parcellation scheme.

### Thalamus Connectivity with 11-Parcellation

Given the relatively stable parcellations observed with the 8-parcellation and the noisiness observed in the 23-parcellation, we experimented with 11 cortical regions that could serve as a middle ground and balance detail with stability/noise. This scheme subdivides the visual cortex into a primary “visual” region and a “visual stream” region, separates the cingulate tract and posterior opercular regions, and introduces an auditory ROI derived from the temporal cortex. The “frontal” region used in the 8-parcellation was reorganized. The results are shown in **Figure 2B** and **Figure 3B**. The resulting 11-parcellation provides a somewhat more fine-grained thalamic map although some limitations persist. Notably, connections with the auditory regions are significantly absent after the winner-takes-all parcellation, perhaps due to significantly poor streamline generation between the thalamus and auditory cortex. Interestingly, connections to the temporal region are observed in the anterior aspect of the thalamus, which aligns with the findings of Behrens (**Figure 2B**, **Figure 2D**).

### Comparative Analysis of Thalamic Parcellation Methods

Comparative axial and coronal views at the same MNI slice level are shown for 8-parcellation CBP with the corresponding ODF clustering, TDI, and THOMAS segmentation and the histologically derived Morel Atlas (**Figure 4**). The methods showed distinct patterns depending on the basic characteristics each method relies on for parcellation. THOMAS leverages the intrinsic contrast contours produced by the WM-nulling of the WMn MP-RAGE sequence and is not surprisingly, closest to the Morel atlas. TDI which uses very short-range fibres and super resolution comes next. ODF also uses diffusion microstructure but is at a lower resolution than TDI and produces clusters which comprise multiple nuclei in some cases (AV+VA for example).

**Figure 4:**
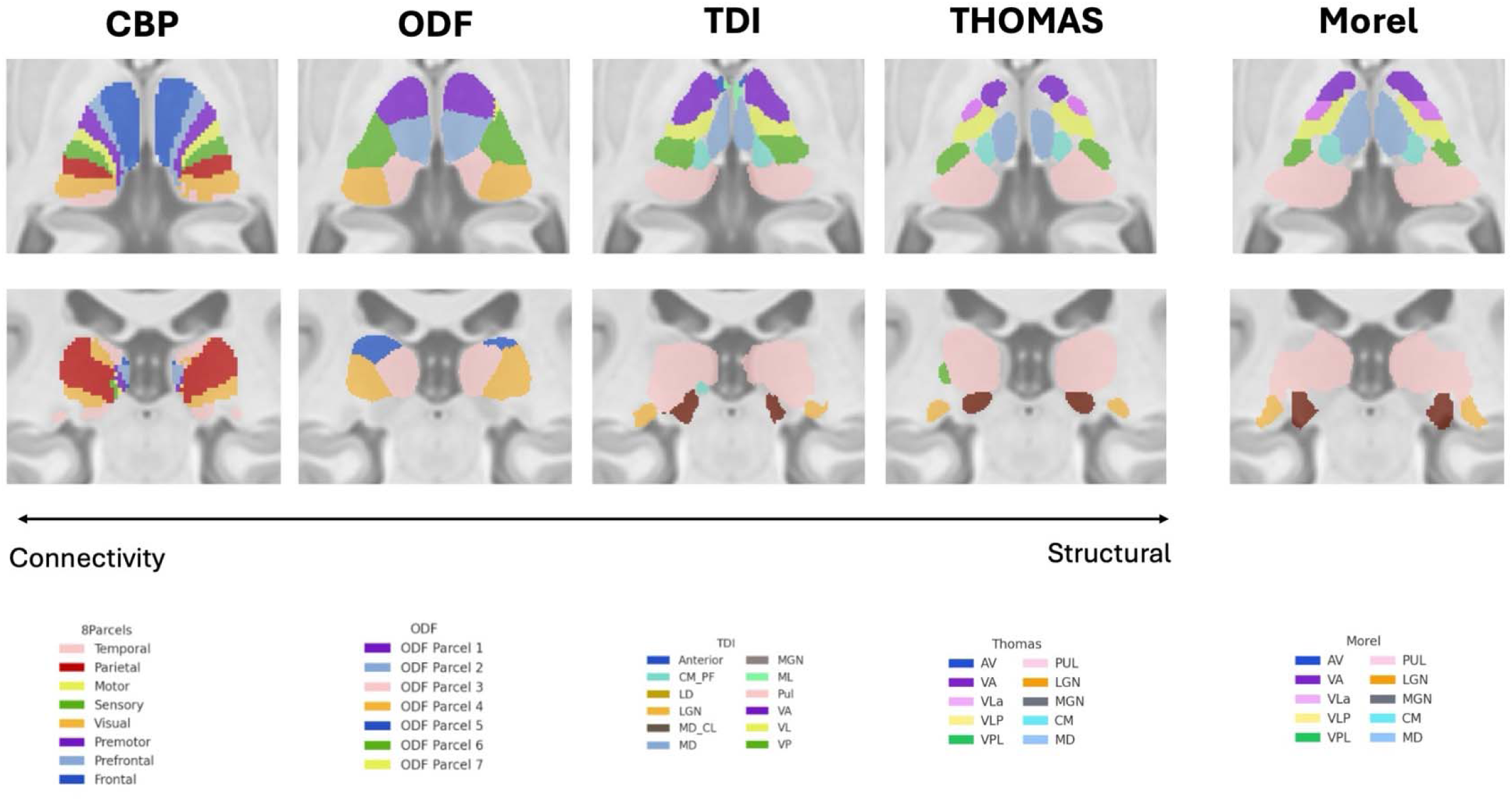
Axial and Coronal View of the 8 Parcellation with Same-Slice Comparisons to ODF, TDI, THOMAS, and Morel Atlas (Relative Maximum Probability) in Axial. The following figure depicts the same axial slice of the MNI template (z = 3) or coronal slice (y = - 27) and the CBP outputs for the 8 regions in comparison to other parcellation methods. ODF clustering of the same n = 67 HCP subjects is also shown. TDI on n = 67 HCP subjects is also shown for comparison. Morel atlas is shown on the far right. THOMAS was run on the same n = 67 HCP subjects, with the outputs shown with relative maximum probability.

Finally, CBP with its long-range connectivity looks farthest from the Morel architectonic parcellation as it is both dependent on functional information (specific cortical regions) and long-range connectivity. Interestingly, the subdivisions of where the pulvinar nucleus are also seen in in both the CBP and ODF clustering, with the lateral parts connecting to visual (occipital) cortex and medial parts connecting to the temporal cortex. For a more quantitative view of the comparisons, Dice overlap scores computed for all 4 parcellation methods (CBP, ODF, TDI, and THOMAS) with the Morel Atlas serving as ground truth is shown in **Figure 5**, where the cluster or parcellation showing the highest Dice is used. (**Figure S3** show a more confusion matrix style 2D plot of each cluster with each nucleus). Highest dice scores were noted in the sensory and frontal parcellations from the 8-parcellation, with dice scores > 0.50 for both (**Figure S3A**). ODF achieved the greatest dice scores for Parcel 2 (dice = 0.659) and Parcel 4 (dice = 0.594), which overlap with the mediodorsal nuclei and the pulvinar nuclei from the Morel atlas (**Figure 4**, **Figure S3B**). TDI resulted in the highest dice scores of 0.550 and 0.603 for the anterior and pulvinar parcellations (**Figure S3C)** Finally, THOMAS resulted in the highest dice scores on average across all parcellations, achieving dice scores of 0.769 and 0.719 for VLP and VPL nucleus, respectively (**Figure S3D)**.

**Figure 5:**
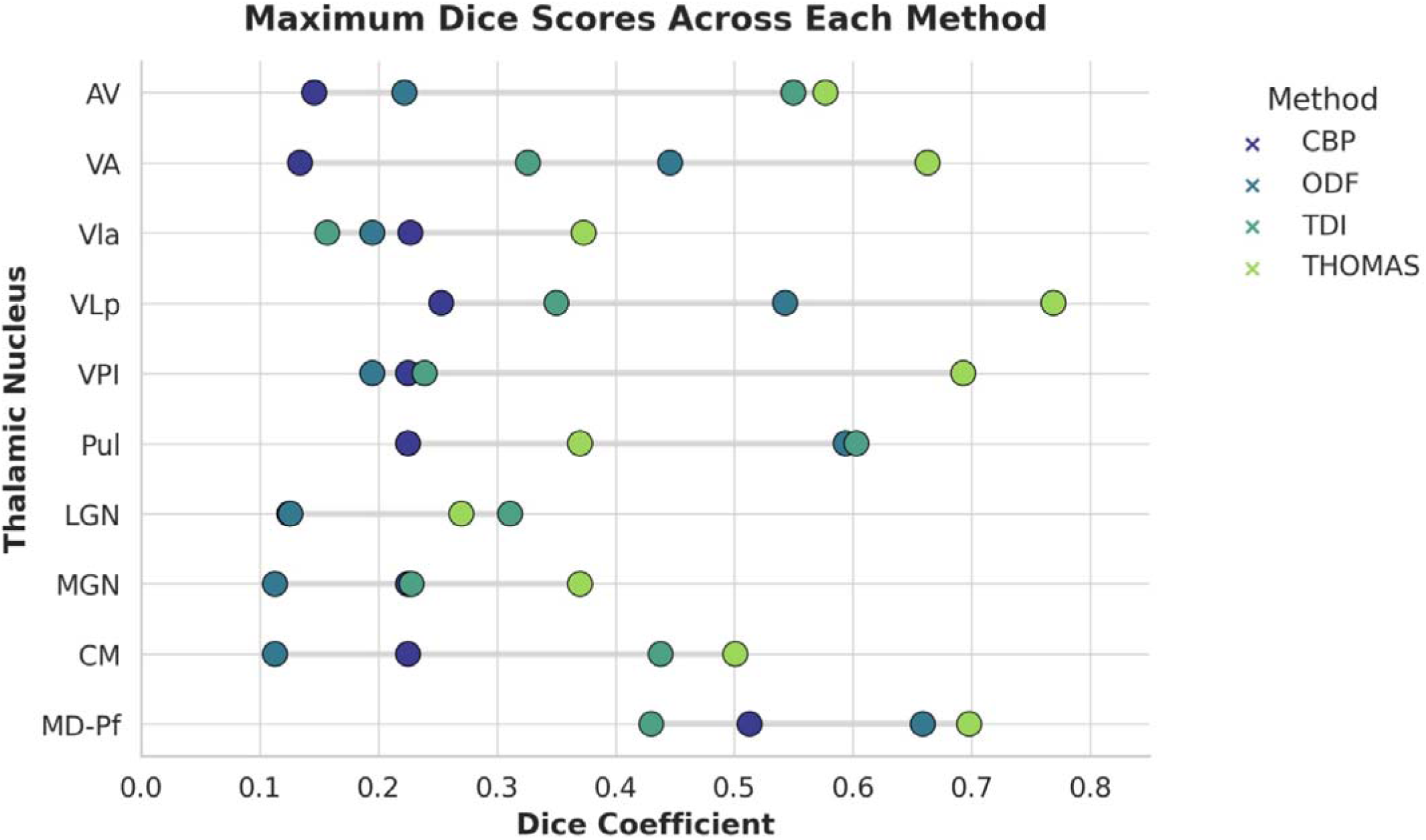
Dice comparison between Morel Atlas versus all 4 methods (CBP, ODF, TDI, Structural). Dice was conducted between each of the parcels in the Morel atlas and compared against each of the parcels in the 8 parcellation, ODF Clustering, TDI, and Structural (THOMAS). Higher dice signify greater overlap between the two parcels.

## Discussion

Reliable and anatomically valid parcellation of the thalamic nuclei remains challenging in both neuroscience and clinical applications. The CBP method was introduced by Behrens et al. in 2003 nearly two decades ago. In this study, we revisited CBP using high-resolution diffusion MRI, sophisticated diffusion MRI reconstruction methods that can handle crossing fibres with facility, and modern cortical parcellations from the HCP-MMP1 atlas. Further, we directly compared CBP results with recent best-in-class segmentation methods: ODF clustering, TDI, and THOMAS, and evaluated their similarities/differences to the Morel cytoarchitectonic atlas. Our results demonstrate that, despite two decades of methodological progress, CBP shows very little changes in anatomical structure relative to its original implementation.

Across our comparative analyses (**Figure 4**) other methodologies (ODF, TDI, THOMAS) exhibited a closer spatial correspondence to the Morel atlas than CBP, particularly in the MD and Pul nuclei. Notably, ODF consistently outperforms CBP in terms of Dice overlap while also resolving the Pul nuclei into subdivisions that mirrored its known lateral (visual) and medial (temporal) connectivity and cytoarchitectonic profiles.^4,8,46^. This finding likely reflects the fact that ODF clustering exploits intrinsic fiber orientation architecture rather than relying on predefined cortical targets, thereby preserving local topography and recovering boundaries that align more closely with cytoarchitectonic maps. TDI also benefits from high spatial resolution and directionality information, producing fine-grained patterns that even further resemble histological maps. THOMAS, however, achieved the highest Dice coefficients on average across all nuclei against all other methods, reaffirming its utility and resourcefulness for anatomical delineation. It should also be noted that the THOMAS framework leverages structural MRI priors that are explicitly derived from the Morel histological atlas, thereby anchoring its segmentations to cytoarchitectonically defined thalamic boundaries. By contrast, CBP frequently produced parcels that did not align well with histological boundaries, and its performance was particularly limited in smaller nuclei.

Several methodological characteristics may contribute to CBP’s persistent limitations. First, the reliance on winner-take-all assignment converts marginal differences in neighboring track density maps into absolute labels, such that even minor fluctuations can flip voxel classification and destabilize parcellation boundaries (resulting in noise). Further, Clayden et al. (2019) have demonstrated that even when internal diffusion orientation information is randomly shuffled, voxel assignments remained consistent, suggesting that CBP parcellations may reflect extrinsic factors such as voxel proximity to major white matter pathways rather than internal thalamic organization ^47^. Our findings also confirm that increasing the number of cortical targets, from 8 to 23 in this study, does not improve boundary definition, but instead exacerbates variability and noise, especially in posterior thalamic nuclei. This increased noise was observed in the 11- parcellation as well. Taken together, this suggests that higher cortical target counts directly contribute to streamline tract variability without adding meaningful anatomical precision. Even with improved diffusion modeling via MSMT-CSD and higher spatial resolution (1.25 mm isotropic compared to the original 3mm isotropic), histological validity appears constrained due to fundamental tractography limitations. However, these results are to be interpreted with caution since the Morel atlas is primarily a structural atlas with minimal functional information included. Its delineations were guided in part by immunohistochemical staining of calcium-binding proteins ^17^ (parvalbumin, calbindin, and calretinin), which reveal differential expression patterns across thalamic nuclei. However, this approach still reflects structural cytoarchitecture rather than specific functional connectivity to cortical regions.

Nonetheless, these results suggest that CBP may be less well suited for applications requiring precise nuclear localization, such as DBS or MRgFUS, where higher spatial accuracy is critical. In the thalamus, our findings indicate that structural methods like THOMAS and diffusion microstructure–based approaches such as ODF clustering and TDI can provide complementary strengths and may represent more reliable options in these contexts. Future work should therefore prioritize hybrid workflows that integrate these strategies, allowing investigators to balance connectivity, cytoarchitectonic, and microstructural information according to the needs of a given study.

In summary, our results indicate that, despite two decades of continued development, CBP retains some of the limitations of its original formulation. While it remains a widely used and valuable tool for broad thalamocortical mapping, our comparisons show that THOMAS achieved the highest overall accuracy relative to histology-based atlases, while ODF clustering and TDI were most effective at delineating anatomically consistent nuclear boundaries. These observations underscore the importance of carefully considering the strengths and limitations of different segmentation strategies, with approaches that directly model local fiber architecture holding promise for bridging in vivo imaging and histological precision.

## Data Availability

All data produced in the present study are available upon reasonable request to the authors

## Acknowledgements

We would like to acknowledge funding from the National Institute of Biomedical Imaging and Bioengineering (R01 EB032674).

## Funding sources

National Institute of Biomedical Imaging and Bioengineering (R01 EB032674).

## Conflicts of Interest

None

## Notes

### Competing Interest Statement

The authors have declared no competing interest.

### Funding Statement

This study was funded by the National Institute of Biomedical Imaging and Bioengineering (R01 EB032674).

### Author Declarations

https://www.humanconnectome.org/study/hcp-young-adult/document/1200-subjects-data-release

