## Supplemental Figures S1, S2, and S3 for "Revisiting the Role of Structural Connectivity-Based Parcellation in Thalamic Nuclei Segmentation: comparison with recent state-of-the-art methods"

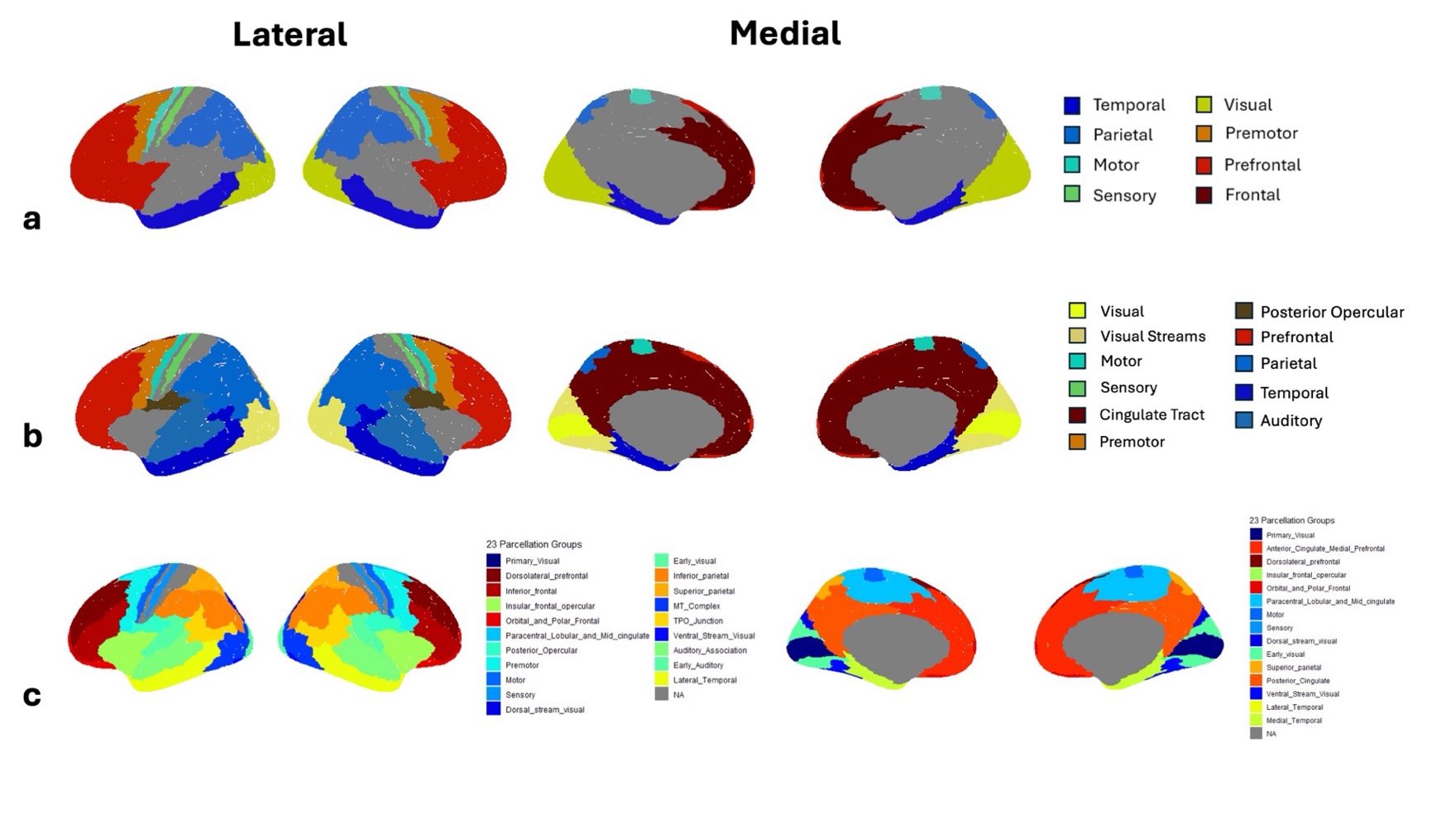

**Figure S1: Cortical Regions defined by the Glasser Atlas used in each CBP Scheme.** The following figure visualizes the cortical regions defined by the 8 parcellation (a), the 11 parcellation (b), and the 23 parcellation (c).

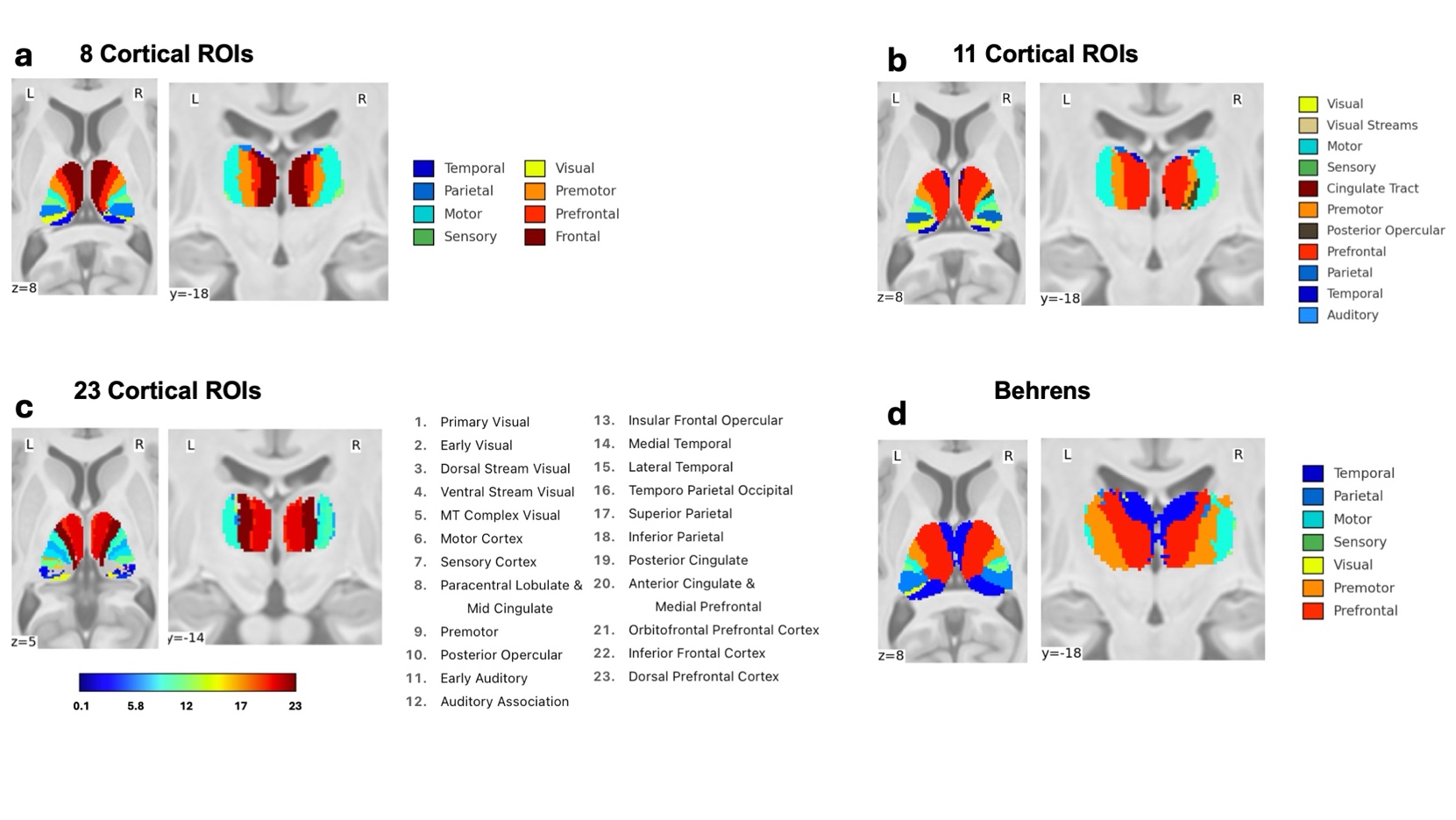

**Figure S2: CBP Between Thalamus and 8/11/23 Cortical Regions of Interest with Comparison to Behrens at 25% Maximum Probability Thresholding.** The following figure depicts cortical-based parcellation (CBP) between the thalamus mask (segmented using THOMAS) and generating streamlines to 8, 11, and 23 cortical regions referenced from Glasser et al, 2016. “8 Cortical ROIs” depicts the 8 parcellation at 25% maximum probability thresholding. “11 Cortical ROIs” depicts the 11 parcellation at 25% maximum probability thresholding. “23 Cortical ROIs” depicts the 23 parcellation at 25% maximum probability thresholding. The Behren parcellation in MNI space is also shown at a 25% maximum probability thresholding for comparison.

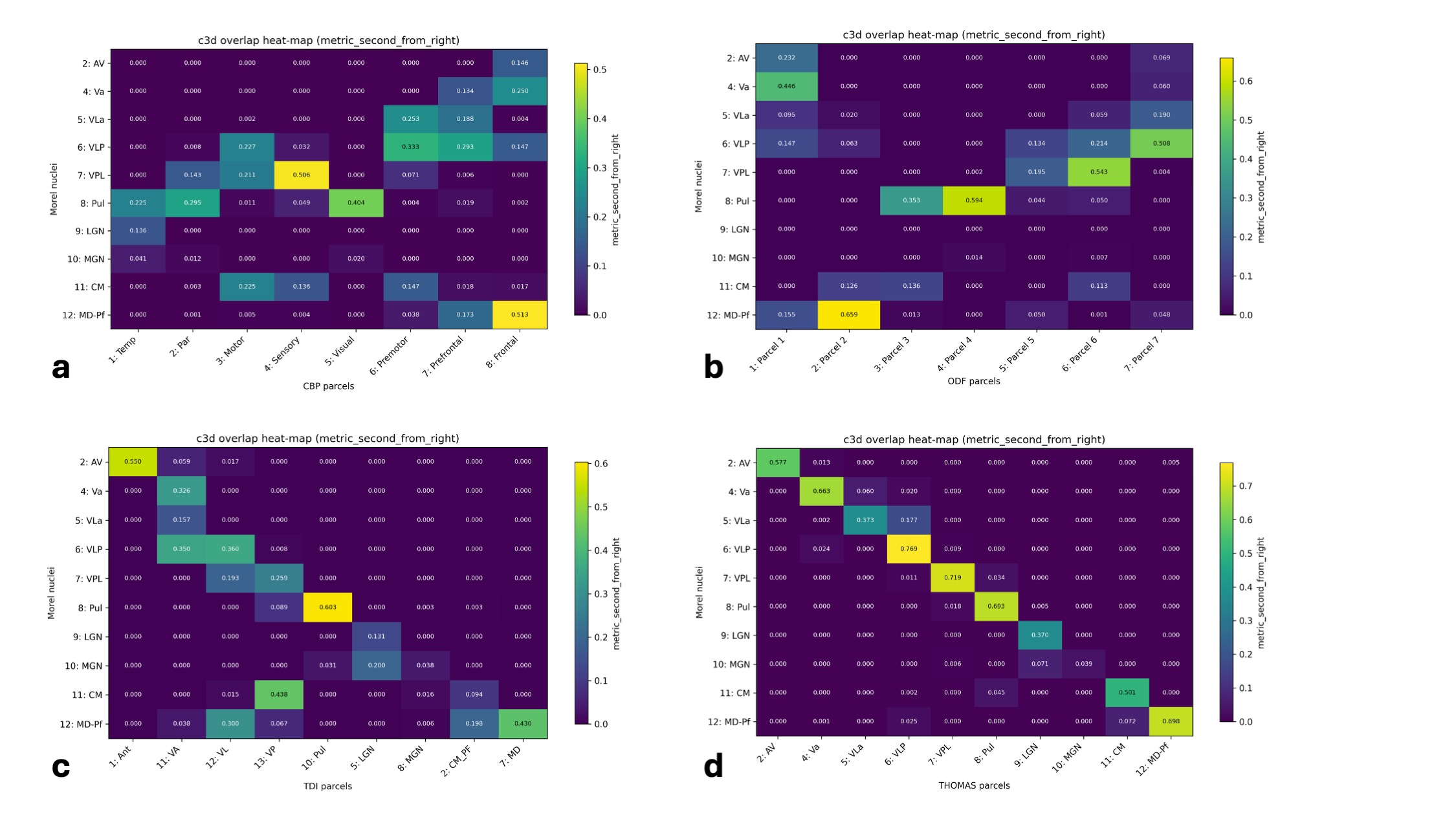

**Figure S3: Dice comparison between Morel Atlas versus all 4 methods.** Dice was conducted between each of the parcels in the Morel atlas and compared against each of the parcels in the 8 parcellation (a), ODF Clustering (b), TDI (c), and Structural (THOMAS) (d). Higher dice signify greater overlap between the two parcels.
